# Barriers to the use of methylphenidate in paediatric neuro-oncology services

**DOI:** 10.1101/2021.12.30.21268574

**Authors:** Alexander J. Hagan, Simon Bailey, Sarah J. Verity

## Abstract

**Background:** The increasing effectiveness of childhood cancer treatment has resulted in a greater number of children surviving previously incurable central nervous system tumours. This growing population of survivors report significant treatment-related difficulties, including attentional impairment associated with poor long-term intellectual development, academic attainment, and health-related quality of life. Clinical findings show benefit to attention and executive functions following methylphenidate administration. The current project explored barriers associated with use of methylphenidate in paediatric neuro-oncology services in the UK.

**Method:** Qualitative data was gathered by semi-structured questionnaire sent to clinical psychologists/neuropsychologists in 19 of the 21 NHS primary treatment oncology centres in the UK in May 2018. Thematic analytic methods were used to explore the data.

**Results:** 11 responses were received from primary treatment centres. Knowledge of the evidence base for methylphenidate in paediatric brain injury was limited. This was primarily attributable to the inadequate resource of psychology into many primary treatment centres, limiting provision to service to a restricted proportion of the patient group. Psychologists reported an interest in exploring the utility of methylphenidate in their patient group. Respondents highlighted the need for provision of accessible research summaries and treatment protocols addressing the potential use of psychostimulants, stating that these would support their team to consider expanding the interventions offered.

**Conclusions:** The development of shared resources for clinicians will be important in supporting the application of research findings to clinical practice. We anticipate national collaboration will support the advancement of intervention for the growing clinical population of long-term survivors.

---

The increasing effectiveness of childhood cancer treatments since the 1970s has resulted in a greater number of young people surviving previously incurable CNS tumours. This growing population of survivors report significant treatment-related difficulties. Both *patient-specific* (e.g. younger age at diagnosis/treatment) and *treatment-specific* (e.g. cranial-directed radiotherapy) factors are associated with neurocognitive deficit. Decline in cognitive processing skills – the ability to take in and make sense of new information - are well documented in survivors of tumour-related hydrocephalus, those treated with cranial radiation therapy, or intrathecal methotrexate (de Ruiter et al., 2013; Robinson et al., 2010). Reduction in processing ability is attributable to the reduced development of white matter in the frontal and parietal lobes following brain injury in childhood. These areas of the brain are developmentally slower to reach full maturation, and thus disproportionately vulnerable to the effects of early injury (Brinkman et al., 2012). The affected rate at which survivors process new information hinders working memory and attentional functioning, consequently lowering the overall level of intellectual ability in the child at maturity.

As the primary neurocognitive deficits reported by survivors of childhood cancer are those mediated by the frontal lobes, psychostimulant medication has been proposed as a rehabilitative strategy (Conklin et al., 2007, 2009, 2010). Clinical findings in paediatric populations show significant increase to attention and executive functions following methylphenidate administration (Nicholls et al., 2012; Smithson et al., 2013). In the United Kingdom, methylphenidate is used as first-line treatment for Attention Deficit Hyperactivity Disorder. To date methylphenidate is prescribed ‘off-label’ for children with acquired attentional deficit secondary to a brain tumour (National Institute for Health and Care Excellence, 2018). Few paediatric neuro-oncology services have formal methylphenidate protocols, and systematic prescribing remains inconsistent.

The current project explored the barriers associated with use of methylphenidate as a potential rehabilitative strategy in paediatric neuro-oncology services in the UK.

## Method

Qualitative data was gathered by semi-structured questionnaire sent to clinical psychologists/neuropsychologists in 19 of the 21 NHS primary treatment oncology centres in the UK in May 2018. The questionnaire targeted current methods of managing treatment-related cognitive impairment, and specifically to the medical management of late effects including attentional deficit. Data were analysed using thematic analysis and accompanied by an iterative categorisation technique (Braun & Clarke, 2006). Independent coder checking was conducted to limit potential bias.

## Results

11 responses were received from the NHS primary treatment centres (58% response rate). Five themes were summarised from the collected data following thematic analysis and organised as below (Figure 1).

**Figure 1.**
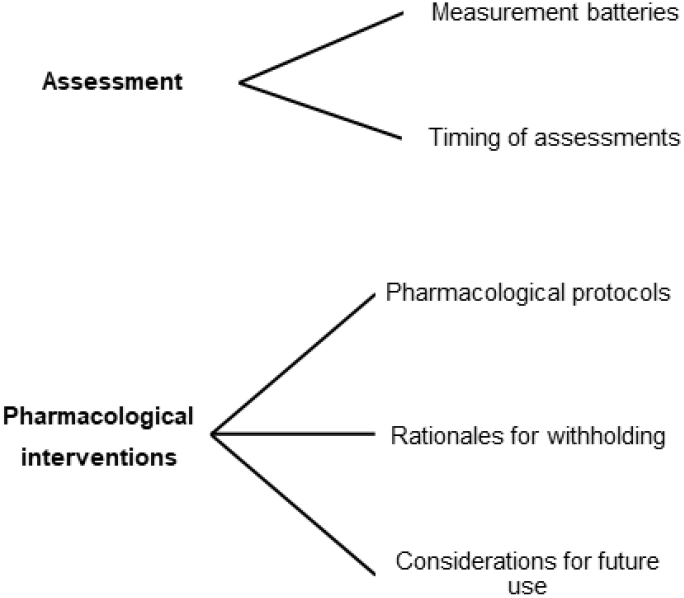
Themes associated with the assessment and pharmacological intervention of attentional impairment.

### Current practice screening attentional impairment

The majority of UK centres employed the same standardised measures to assess neurocognitive difficulties in survivors of childhood cancer. Wechsler Intelligence Scales (e.g. WISC-V / WPPSI-IV) and their appropriate timed subtests (e.g. Coding / Bug Search) were used within NHS centres as a measure of processing speed. Attentional functioning, where assessed, was predominately measured using a version of the Test of Everyday Attention for Children (TEACh or TEACH 2), or in two centres by the Conners’ Continuous Performance Test. Attentional function specifically was not commonly screened as routine practice in the majority of centres.

Most centres assessed survivors at six months to one-year following treatment involving cranial radiation therapy. There was significantly less frequent provision of routine screening to survivors whose treatment included surgery only, or chemotherapy only. Routine long-term follow-up screening of neurocognitive functioning also varied substantially, and depended on the capacity and resource of qualified clinical psychologists/ neuropsychologists. Resource limitations frequently guided the provision and timing of screening assessments:

*‘Honestly not often enough! When we have to, when we can, when people are worried. We are trying to move to a pathway and regular assessment time points but we are just not sufficiently resourced for this at present*.*’*

### Current use of psychostimulant medication for management of attentional deficit in paediatric neuro-oncology survivors

Psychostimulants were rarely used for managing the neurocognitive deficits identified in survivors of childhood cancer. Of the 11 centres that responded, only one centre prescribed psychostimulants routinely as part of a rehabilitative strategy managed by the neuro-oncology MDT. Some centres reported the occasional use of psychostimulants, but solely when comorbid diagnoses were present:

*‘No – unless, persistent post-traumatic fatigue is seen of ADHD/ADD is suspected’*

In centres in which psychostimulants were on occasion prescribed, psychiatry was the discipline responsible for this.

### Rationales for not using pharmacological strategies

One of the main emerging barriers to the use of psychostimulant rehabilitative strategies was due to a perceived lack of evidence:

*‘I think that the reason why it is not offered routinely is because the limited number of research studies available that highlights the benefits*.*’*

*‘There is limited evidence to support the use of pharmacological approaches …’*

A barrier expressed by a number of respondents was distinguishing a colleague within their multi-disciplinary teams who would monitor psychostimulant interventions.:

*‘There can be variation in familiarity and willingness among psychiatry and paediatric colleagues to prescribe medicines for cognitive prob[lem]s…’*

Many responses indicated a perceived belief that a paediatric psychiatrist would be required to support prescribing of psychostimulants:

*‘…we have limited psychiatry time so regular prescription is difficult*.*’ ‘We don’t have a specialist in this area…’*

Two respondents raised concerns regarding the potential side effect profile of psychostimulants in survivors of childhood cancer. The perceived low acceptability of further medical intervention to parents of survivors was also cited as a barrier to considering this treatment option:

*‘I assume the usual stimulant side effects profile might cause concern for CYP [Children and Young People], family, or medical teams*…*’*

*‘… [Parents would be] concerned about their child taking medication, as they will already have had a number of potential side effects from chemo[therapy]’*

### Interest in potential use of psychostimulant treatment

The majority of respondents reported an interest in the potential utility of psychostimulants in paediatric neuro-oncology survivors, and stated a wish for further evidence demonstrating the efficacy of psychostimulants in this population. Clinicians reported being aware of relevant published literature, but believed that this provided insufficient evidence to inform their own practice:

*‘If the evidence-base was sufficiently robust, then I am confident our team would use this to inform practice*.*’*

*‘We don’t have the time to stay ahead of emerging research – we need protocols and clinical summaries’*

Whilst resource barriers limited the access of some teams to pharmacological interventions, the majority of respondents were keen to explore their potentially utility:

*‘The team is always interested in new techniques to improve outcomes for our patients’ ‘I’m definitely interested in trying this’*

## Discussion

Survivors of childhood cancer are at risk of significant neurocognitive late effects. The potential utility of psychostimulants in partially alleviating these difficulties is reported in a number of clinical trials. Nonetheless, the use of psychostimulants for survivors of childhood cancer depends on the preference or knowledge base of individual clinicians, and the resource of individual centres to provide appropriate assessment and management of the treatment. The present project explored the use of psychostimulant medication as part of a rehabilitation strategy in specialist neuro-oncology centres in the UK.

We found knowledge of the existing literature base for psychostimulant use in this population to be limited in many centres. Despite the growing evidence base for the utility of psychostimulant medication in children with acquired brain injury via traumatic brain injury or tumour treatment, use of methylphenidate is not considered as a routine option in mainstream clinical practice. Respondents noted that the highly-constrained resource available to paediatric psychologists in many specialist treatment centres restricts practice in many cases to provision of a skeleton service. This means that only the most clinically necessary cases are assessed, and even fewer supported with rehabilitation. In such an environment, there is little time available for the consideration of emerging interventions, even less scope to engage in research of novel therapeutic strategies or intervention. This remains an ongoing source of frustration to many clinicians, whose doctoral training as research clinicians is unsupported and under-utilised in the poorly-resourced environment of NHS clinical practice.

Despite such limitations, we found psychologists in the specialist treatment centres to be interested in supporting pharmacological interventions, and keen to gain a greater understanding of the potential for these. Respondents highlighted the need for the provision of accessible research summaries and treatment protocols addressing the potential use of psychostimulants, stating that these would support their team to consider expanding the interventions offered.

We have offered a ‘Survivorship clinic’ at the Great North Children’s Hospital in Newcastle since April 2017. This co-led clinic is supported by a paediatric neuropsychologist, a consultant paediatric neuro-oncologist, and a specialist nurse practitioner. It is held approximately twice per month, providing screening, assessment, and methylphenidate intervention where appropriate to paediatric neuro oncology survivors. We have found significant benefit to our patients via this intervention, and are in the process of preparing a UK wide four-centre study to support our data. Our clinical information sheets for parents and children, and our assessment, prescribing, and follow-up protocols are available on request by any UK centre. We anticipate that national collaboration will support the advancement of intervention for the growing and important clinical population of long-term survivors.

## Data Availability

All data produced in the present study are available upon reasonable request to the authors

## Acknowledgements

Tiara Pratomo (School of Medicine, Newcastle University) assisted with data collection as part of her own MRes. Uschi Hiermeier (Assistant Psychologist, Newcastle Upon Tyne Hospitals NHS Foundation Trust) assisted with manuscript preparation.

